# A reliable and valid measure of COVID-19 patient-reported symptoms in outpatients: the Symptoms Evolution of COVID-19 (SE-C19) instrument

**DOI:** 10.1101/2021.12.16.21267708

**Authors:** Diana Rofail, Pip Griffiths, Giulio Flore, Mohamed Hussein, Sumathi Sivapalasingam, Anna J. Podolanczuk, Ana Maria Rodriguez, Vera Mastey, Chad Gwaltney

**Affiliations:** Regeneron Pharmaceuticals Inc., Sleepy Hollow, NY, USA; IQVIA, Cambridge, MA, USA; Weill Cornell Medical College, New York, NY, USA; Gwaltney Consulting, Westerly, RI, USA

## Abstract

**Background:** There is no valid and reliable patient self-reported measure assessing symptomology among outpatients with COVID-19. The Symptoms Evolution of COVID-19 (SE-C19) is a self-administered new instrument that includes 23 symptoms, each rated for severity at their worst moment within the last 24 hours. We studied the psychometric properties of SE-C19.

**Methods:** Reliability, validity, and sensitivity to change of the SE-C19 were assessed in 657 outpatients with confirmed COVID-19 enrolled in NCT04425629. SE-C19 and Patient Global Impression of Severity (PGIS) were administered daily from baseline (predose at Day 1) to end of study (Day 29).

**Findings:** Most patients (70·0%) were aged ≤50 years and white (85·5%). At baseline, patients reported an average (SD) of 6·6 (3·9) symptoms (ie, rated as at least *Mild)* with 3·8 (3·3) of these symptoms being rated as *Moderate or Severe*. By Day 29, most symptoms had resolved; 74·4% of patients reported no symptoms and on average, only 0·6 (SD 1·5) symptoms were reported as at least *Mild*.

Stable patients according to the PGIS showed scores with intraclass correlation values indicating moderate-to-good test-retest reliability (ie, 0·50-0·90). At baseline, 20 item scores (87%) varied significantly across PGIS defined groups supporting the validity of SE-C19.

A symptom-resolution endpoint was defined after excluding the item “Sneezing”, due to its low ability to discriminate severity levels, and “Confusion”, “Rash”, and “Vomiting”, due to their low prevalence in this population. Symptoms resolution required complete absence of all remaining items, except “Cough”, “Fatigue”, and “Headache”, which could be *Mild* or *Moderate* in severity.

**Interpretation:** We identified 19 items that are valid and reliable to measure disease-related symptoms in COVID-19 outpatients and propose a definition of symptom resolution that could be used in future clinical trials and potentially, also in clinical practice.

**Funding:** This research was funded by Regeneron Pharmaceuticals, Inc.

**Research in context:** *Evidence before this study:* The majority of COVID-19-infected patients (>80%) have mild-to-moderate symptoms and are managed at an outpatient setting. Although clinical research has primarily focused on prevention of the disease and the treatment of hospitalised patients, close monitoring of the COVID-19 symptoms and their severity in outpatients is equally important and needed to prevent community transmission. Patient-reported outcome (PRO) instruments are a key method to assess COVID-19 related symptoms and associated burden as these symptoms are best known by the patient and are best measured from the patient perspective. However, a valid and reliable instrument to assess symptom severity and progression among outpatients with COVID-19 is not yet available. This study focuses on the psychometric properties (reliability, validity, and sensitivity to change) of a new recently developed self-administered PRO symptom measure (SE-C19) for COVID-19-positive outpatients.

*Added value of this study:* This is the first study to systematically examine the psychometric properties of a PRO symptom measure designed for COVID-19 outpatients and it provides a method for identifying symptom resolution among outpatients, which may be useful in clinical research and clinical practice contexts. The SE-C19 instrument is a self-administered questionnaire that assesses the severity of 23 COVID-19-related symptoms. The recall period is 24 hours and the response options include *None, Mild, Moderate* and *Severe*. The analyses reported here demonstrate that the SE-C19 is a valid and reliable measure to capture daily COVID-19 symptom severity from the outpatient’s perspective. These psychometric analyses also provide empirical evidence for a method to determine symptoms resolution based on the score of 19 of the SE-19 items; this may be useful not only in clinical trials but also in real-world studies and clinical practice. The 4 items not included in the symptoms resolution endpoint may be useful to clinicians to monitor severe disease. The SE-C19 instrument is relevant for the clinical management of outpatients, as it measures a large number of symptoms that are relevant in the outpatient setting and reflect the heterogeneity of symptom experience, and it is sensitive to the longitudinal changes in the severity of these symptoms.

*Implications of all the available evidence:* The SE-C19 can be used to monitor COVID-19-related symptoms over time in outpatients. The definition of symptoms resolution established here can be used to inform clinical trial endpoints and may also be useful in clinical practice to aid discussions between healthcare professionals and patients, and inform treatment decisions. Symptoms resolution is based on 19 of the 23 items included in the SE-C19 instrument.

## Introduction

Clinical presentation of the novel coronavirus disease 19 (COVID-19) is heterogenous, ranging from patients being asymptomatic to having severe disease manifestations and life-threatening complications that require hospitalisation and intensive care.^1^ The prevalence of asymptomatic COVID-19-infected patients is not yet well understood; rates depend on the extent of testing and may also depend on the prevalent COVID-19 variant(s) in each geographic region.^2^ In addition, many patients who are asymptomatic at diagnosis may develop symptoms at a later stage (ie, they are presymptomatic).^3^

Much of the clinical research has focused on developing vaccines to prevent COVID-19 infection and new therapies for severe and/or hospitalised COVID-19 patients. However, most symptomatic patients (>80%) have mild-to-moderate symptoms and are being managed by primary healthcare providers.^4^ This percentage is expected to increase as COVID-19 vaccination coverage gradually expands globally and patients develop less severe disease.

COVID-19 affects different people in different ways. A wide range of COVID-19 symptoms encompassing several organs has been reported by outpatients across different studies. Although the symptoms reported by outpatients differ across studies, the most commonly reported symptoms in outpatients include chills, fever, headache, stuffy or runny nose, sore throat, shortness of breath, cough, fatigue, muscle or body aches, nausea, vomiting, loss of smell and loss of taste.^5–11^

When present, COVID-19 symptoms can last days, weeks or even months^1,12^ with symptoms not initially present often being manifested later during the disease. In most patients, symptoms resolve spontaneously however the resolution kinetics differ across symptoms and some symptoms may remain long after viral clearance (ie, post-acute sequelae severe acute respiratory coronavirus 2 [SARS-CoV-2] infection also known as long COVID-19).^12^

Most measures to monitor symptoms evolution have been developed for hospitalised patients and are based on objective measures (eg, monitoring oxygen saturation or haemodynamics), but these measures are rarely applicable to outpatients. Outpatients need to be closely monitored to ensure that symptoms gradually resolve reducing the symptom burden and avoiding severe disease and hospitalisation. Evaluating the longitudinal trajectory of symptoms occurrence and severity requires novel reliable and valid measures tailored to this population. Patient-reported outcomes (PRO) measures are ideal, because they directly reflect the patient’s perspective.^5,13^ In most countries, progression of symptoms in outpatients is often monitored by telematic or telephone follow-up and resolution determined after 10 days from onset of symptoms if absence of fever for at least 3 days and clinical improvement of symptoms other than fever, or if there are two consecutive negative SARS-CoV-2 reverse transcription quantitative polymerase chain reaction (RT-qPCR) tests from respiratory specimens in a 24-hour interval.^14^ A more specific symptom resolution definition is needed to assess the treatment benefit of new COVID-19 therapies for outpatients, in a clinical trial context.

Many of the COVID-19-related symptoms (eg, dyspnoea or chest pain) pose an important burden to patients and are often the main reason for seeking medical care at outpatients or emergency department. Given the symptomatic burden of COVID-19 and the Food and Drug Administration (FDA)’s effort to implement the patient perspective in regulatory decision-making, the FDA has provided guidance to evaluate the burden of COVID-19 in outpatients participating in clinical trials by using PRO instruments that assess a set of COVID-19-related symptoms at least every 24 hours.^5^ Despite this FDA guidance, no valid and reliable instrument to assess outpatient-reported symptom progression in clinical trials or clinical practice is yet available.

Regeneron Pharmaceuticals, Inc. recently developed the Symptoms Evolution of COVID-19 (SE-C19) instrument to address the symptoms experienced by COVID-19 outpatients.^15^ The aim of this study was to assess the extent to which the SE-C19 instrument is valid, reliable, and able to detect symptoms changes in outpatients with laboratory-confirmed COVID-19, and to establish a definition of symptom resolution.

## Materials and methods

### Study population

Psychometric properties of SE-C19 were studied in symptomatic patients recruited in the NCT04425629 (COV-2067) trial, an ongoing, adaptive, phase 1/2/3, randomised, double-blinded, placebo-controlled study assessing the efficacy and safety of the combination of the casirivimab with imdevimab monoclonal antibodies, in adult outpatients with COVID-19.^16^ Patients could be symptomatic or asymptomatic at baseline but all patients needed a positive RT-qPCR in nasopharyngeal swab samples at randomisation. In addition, patients could not have been previously hospitalised nor be currently hospitalised for COVID-19. The study lasted 29 days from randomisation at Day 1.^16^

### Patient-reported outcomes in NCT04425629 (COV-2067)

The PROs administered in the study were SE-C19, the Patient Global Impression of Severity (PGIS), and the Patient Global Impression of Change (PGIC). The two last questionnaires were used as anchors in the assessment of SE-C19 psychometrics. The SE-C19 was developed by Regeneron based on the Centers for Disease Control and Prevention (CDC) symptom list^6^ and available published literature specific to patients with COVID-19 as of spring 2020,^15^ describing the symptomatic course of COVID-19 infection over time. Its development followed the measurement principles outlined in the FDA PRO Guidance,^17^ and the methodological Patient-Focused Drug Development Guidance Series.^18^ Its content validity was confirmed in semi-structured qualitative interviews with COVID-19-positive patients.^15^

The SE-C19 consists of 23 symptoms associated with COVID-19, including respiratory, gastrointestinal, and febrile symptoms. It is administered electronically with patients using their own devices to complete it (i.e., a “bring your own device” approach, or BYOD). If BYOD is not feasible, a web-link is provided to the patient.^15^ Patients are first asked to select all experienced symptoms and are then moved to the next screen where they need to confirm these symptoms and rate the severity of each experienced symptom as *Mild, Moderate*, or *Severe* at its worst moment within the last 24 hours. Symptoms not checked in the initial screen are automatically rated as *None* for that timepoint.^15^ Before completing the questionnaire, patients are given the opportunity to review the selected list of symptoms and associated severity. The SE-C19 was completed daily for a period of 29 days.

At each time point, after completing the SE-C19, patients were also asked to complete the PGIS, a single question that rates the overall symptom severity at their worst moment in the last 24 hours from 0 (*No symptoms*) up to 3 (*Severe symptoms*).^19^ Additionally, at Day 29, patients completed the PGIC to assess patient’s perception of the degree to which their symptoms had changed since baseline on a seven-point scale from -3 (*Very much worse*) to 3 (*Very much better*).^20^

### Statistical analysis

All psychometric analyses were performed on the pooled treatment arms modified full analysis set (mFAS) defined as all randomised patients with a positive RT-qPCR in the nasopharyngeal swab samples at randomisation in NCT04425629 (COV-2067). The analyses were conducted at the individual as well as combined item level.

The distribution of SE-C19 items and PGIS responses were described from baseline up to Day 29 daily, using frequency and percentage for categorical variables and means and standard deviations for continuous variables. These descriptive statistics were used to observe which symptoms were reported at each day and to investigate temporal trends.

Correlations between each individual item were assessed using Spearman correlation coefficients with correlations above 0·4 indicating items, which may form a sub scale or domain.

Test-retest reliability was assessed using intra-class correlation coefficients (ICC).^21^ ICC was calculated for each symptom between pairs of consecutive days from Day 16 to Day 22 in selected “stable” patients (measured by patients presenting the same PGIS score at each pair of days). ICC values of 0·50–0·75 indicated moderate reliability, between 0·75–0·90 good reliability, and values >0·90 excellent reliability.^22^

Ability of the SE-C19 to discriminate among groups of patients (Known-groups validity) was assessed by comparing the difference in SE-C19 score between patients grouped by their self-reported severity at baseline (baseline PGIS score, grouped *none/mild* or *moderate/severe*), using a Mann-Whitney U test.

The ability of the SE-C19 to detect change in disease severity was also assessed. Change in individual SE-C19 symptom scores between baseline and Day 15, and between baseline and Day 29 was assessed for patients who had reported a change in severity at both time periods on PGIS, using a Mann-Whitney U test.^23^ Days 15 and 29 were selected because a change either as a result of the study treatment or to the normal evolution of the condition could be expected at these two timepoints.

Structural validity was assessed using the longitudinal item response theory (IRT), which measures the ability of each item to discriminate across symptoms severities across timepoints. In addition, single-timepoint Rasch Measurement Theory (RMT) models were built based on the results arising from the longitudinal IRT. Iterations continued until model properties were acceptable as defined by the following premises.^24^ First, the response options for each item were appropriate, ie, the ordering of response options was consistently discriminated by patients across the four options (ie, *None, Mild, Moderate*, and *Severe*). Secondly, the range of expected item scores matched the range of expected patient severities on the same measurement continuum, using Wright maps. Thirdly, no additional relationship between items existed beyond those expected by the model (once underlying patient severity has been accounted for). The G2 chi-square statistic was then used to assess the degree of relationship between observed responses after controlling for the underlying severity and tested through the residual inter-item correlations using the Yen’s Q3 statistics. Finally, the overall item fit was assessed using an iterative procedure where fit to the model was assessed both at the item- and patient-level, using “infit” and “outfit” statistics. A rating scale model^25^ was used and adjustments were made where necessary to improve the fit of the SE-C19 to the model.

## Results

### Baseline characteristics

The mFAS criteria was met by 657 patients. Most patients were aged 50 years or younger (70·5%), female (52·1%), white (85·5%) and not Hispanic or Latino (51·3%; Table 1). Overall, 61·6% of patients had one or more risk factors for severe COVID-19 with cardiovascular disease (20·8%) and chronic metabolic disease (13·5%) being the most prevalent (Table 1).

**Table 1:** Demographics and baseline characteristics of mFAS.

|  |  | n (%) |
| --- | --- | --- |
| Total population |  | 657 |
| Population breakdown |  |  |
| Sex | Female | 342 (52.1%) |
|  | Male | 315 (47.9%) |
| Age group | Age ≤50 | 463 (70.5%) |
|  | Age >50 | 194 (29.5%) |
| Ethnicity | Not Hispanic or Latino | 337 (51.3%) |
|  | Hispanic or Latino | 318 (48.4%) |
|  | Not reported | 2 (0.3%) |
| Race | White | 562 (85.5%) |
|  | Black or African American | 61 (9.3%) |
|  | Asian | 9 (1.4%) |
|  | American Indian or Alaska Native | 4 (0.6%) |
|  | Unknown | 5 (0.8%) |
|  | Not reported | 16 (2.4%) |
| Comorbidities | Cardiovascular disease | 137 (20.8%) |
|  | Chronic lung disease | 65 (9.9%) |
|  | Chronic metabolic disease | 89 (13.5%) |
|  | Chronic kidney disease | 10 (1.5%) |
|  | Chronic liver disease | 3 (0.5%) |
|  | Immunosuppressed | 11 (1.7%) |
|  | Taking immunosuppressants | 6 (0.9%) |
|  | Not reported | 2 (0.3%) |
| Risk factor* | No risk factors for hospitalisation due to COVID-19 | 252 (38.4%) |
|  | ≥1 risk factor for hospitalisation due to COVID-19 | 405 (61.6%) |

\*Having a comorbidity is a risk factor for hospitalisation. mFAS=modified full analysis set.

### Patient-reported symptoms at baseline

Completion rates of SE-C19 and PGIS were high at baseline (SE-C19: 83·7%; PGIS: 69·5%) and remained higher than 68% up to Day 22 (Table S1). Thereafter, completion rates fell below 50%. At baseline most patients (532 of the 543 patients completing SE-C19; 98.0%) reported one or more symptoms. On average, patients reported 6·6 (SD: 3·9; median 6·0) different symptoms (i.e., rated as *Mild, Moderate* or *Severe*). Of these, 3·8 symptoms (SD: 3·3; median 3·0) were rated as *Moderate or Severe*, and 0·8 (SD: 1·5; median 0·0) as *Severe*. By Day 29, most symptoms had resolved with only 0·6 (SD: 1·5; median 0·0) symptoms being rated *Mild* or greater (Table 2). Overall, 74·4% (n=238) of patients reported no symptoms at Day 29.

**Table 2:** The number of symptoms at different rating levels at baseline, Day 7, Day 15, Day 22 and Day 29 after randomisation.

| Symptom severity | Baseline | Day 7 | Day 15 | Day 22 | Day 29 |
| --- | --- | --- | --- | --- | --- |
| Number of patients | 543 | 519 | 462 | 488 | 320 |
| At least *Mild* (rating > 0), mean (SD) [median] | 6.6 (3.9)<br>[6.0] | 3.1 (2.9)<br>[2.0] | 1.5 (2.2)<br>[1.0] | 0.9 (1.7)<br>[0.0] | 0.6 (1.5)<br>[0.0] |
| At least *Moderate* (rating > 1), mean (SD) [median] | 3.8 (3.3)<br>[3.0] | 1.2 (2.0)<br>[0.0] | 0.5 (1.4)<br>[0.0] | 0.3 (1.0)<br>[0.0] | 0.2 (0.7)<br>[0.0] |
| *Severe* (rating =3), mean (SD) [median] | 0.8 (1.5)<br>[0.0] | 0.2 (0.6)<br>[0.0] | 0.0 (0.4)<br>[0.0] | 0.0 (0.3)<br>[0.0] | 0.0 (0.1)<br>[0.0] |

SD, standard deviation.

For most symptoms, the proportion of patients reporting not experiencing the symptom (ie, rated as *None*) at baseline (30·9%-97·4%) outnumbered patients reporting it (Figure 1). The 5 most prevalent symptoms at baseline were “Cough” (reported by 69·1% of patients and 4·4% rated it as *Severe*), “Fatigue” (61·1% [*Severe*: 12·0%]), “Headache” (60·6% [*Severe*: 9·4%]), “Body aches” (52% [*Severe*: 10·9%]) and “Loss of taste/smell” (47% [*Severe*: 20·1%]). In the PGIS, most patients scored their COVID-19 symptoms as *Mild* (42·6%) or *Moderate* (47·9%) at baseline with 3·5% and 6·0% of patients reporting *None* and *Severe*, respectively (data not shown).

**Figure 1:**
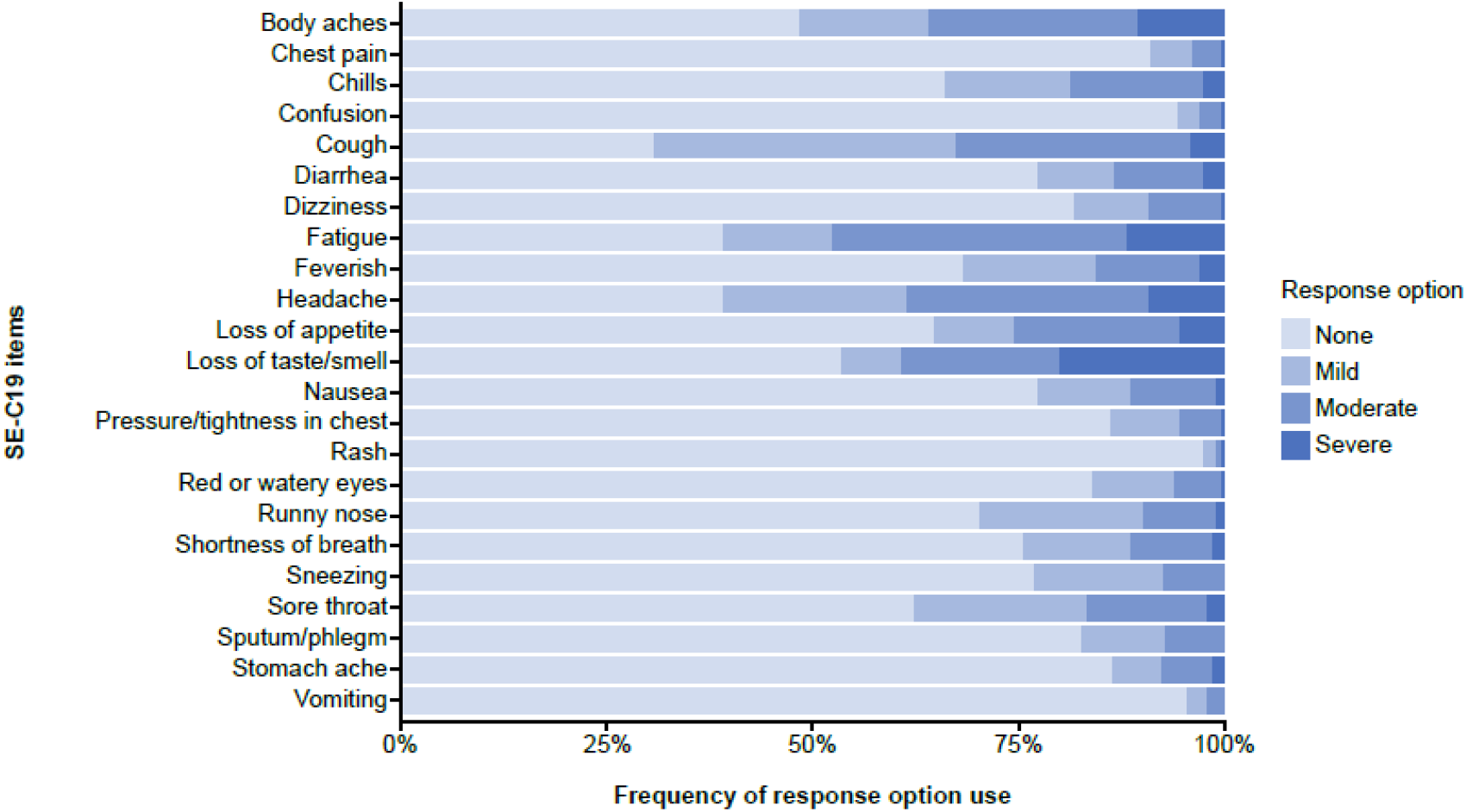
Distribution of responses for SE-C19 items at baseline. SE-C19=Symptoms Evolution of COVID-19.

Symptoms such as “Feverish”, “Vomiting”, “Confusion”, and “Rash” reached a symptom resolution rate above 95% within the first week from randomisation (Table S2). For many other symptoms such as “Chills”, “Nausea”, “Diarrhoea”, “Red or watery eyes”, “Dizziness”, “Pressure/tightness in chest” and “Stomach ache”, a similar resolution rate was not reached until the end of the second week from randomisation. By Day 22, ≥90% of patients reported having “no symptoms” (ie, *None*) for the majority of the items, with the exception of “Cough”, “Headache”, “Loss of taste/smell”, and “Fatigue”, for which symptom resolution rates ranged between 85% and 90%.

The correlation between pairs of items were weak for the majority of cases (r<0·3); 14 items pairs out of 253 had “moderate” correlations (between r=0·3 and r=0·5; Figure S1). The two symptoms pairs that most strongly correlated were “Headache” with “Body aches” (r=0·40) and “Feverish” with “Chills” (r=0·48; Figure S1). Despite some item pairs showing higher correlations, the observed low to moderate correlations did not support the need for specific symptom clusters or overall domain or total scores.

### Reliability, Known-group validity and ability of the SE-C19 to detect change

The test-retest reliability, ie, the ability of the SE-C19 items to provide consistent scores when the symptoms were stable over time, was moderate to good across most items, with ICC estimates ranging from 0·50 to 0·90 (Table S3). However, there were some exceptions. Test-retest reliability could not be tested for “Rash”, “Chest pain”, “Confusion”, or “Vomiting” because of low prevalence at the assessed time periods. In addition, “Red or watery eyes”, “Stomach ache”, and “Dizziness” had varying profiles of reliability, where acceptable results were seen for some time periods but not for others.

Patients with a higher self-reported COVID-19 symptom severity, as measured by the PGIS at baseline, reported more severe mean SE-C19 item scores (Table S4). Out of the 23 items, 20 items (87%) had a statistically significant difference in SE-C19 scores between the PGIS severity groups. The magnitude of differences varied across items, with “Fatigue” showing the largest difference between groups (mean difference=0·72 out of a maximum possible difference of 4), and “Rash” showing the smallest difference (mean difference=0·04 out of a maximum possible difference of 4).

Most SE-C19 items were able to detect change with scores decreasing over time among patients who reported improvement on the PGIS (Table S5) or PGIC (Table S6). The exceptions were the items “Confusion”, “Rash”, and “Red or watery eyes”, for which the direction of the score change on SE-C19 was misaligned with the reported improvement on the PGIS or PGIC. The ability of each item to discriminate between symptoms severities, using longitudinal IRT, supported that “Sneezing” had poor discriminatory ability and therefore was excluded from the RMT analyses.

Examination of the category probability curves using the RMT analyses suggested that patients were unable to reliably distinguish option *Mild* from *Moderate* (Figure 2). As a result, these two response options were merged into a single response score representing “*Mild/Moderate*” and the RMT model rerun. In the new model, “Vomiting”, “Confusion”, and “Rash” fell outside of the expected severity range for this study population with *Mild*/*Moderate* rating being associated with a latent score >3 suggesting that very few patients would report these symptoms (see Figure 3). When assessing whether high correlations between items existed beyond those expected by the model (ie, once the underlying patient severity has been accounted for), most items had a low correlation level (<0·20). “Chills” and “Feverish” had the highest residual correlation of 0·31.

**Figure 2:**
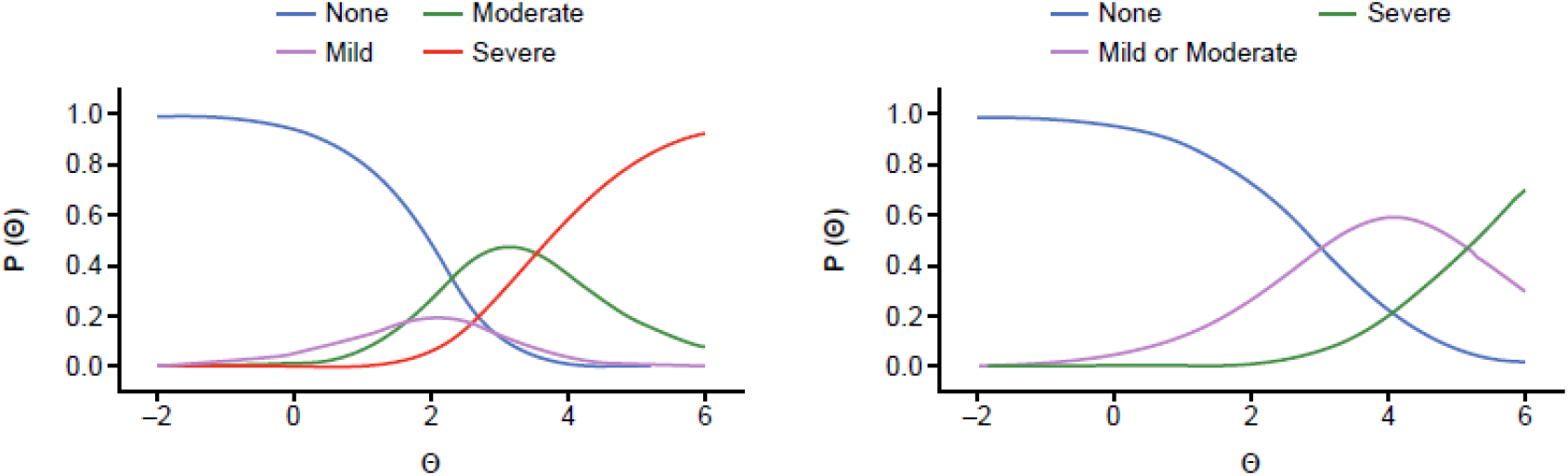
Example of item response curves when using all four response options compared to merging *Mild* and *Moderate*.

**Figure 3:**
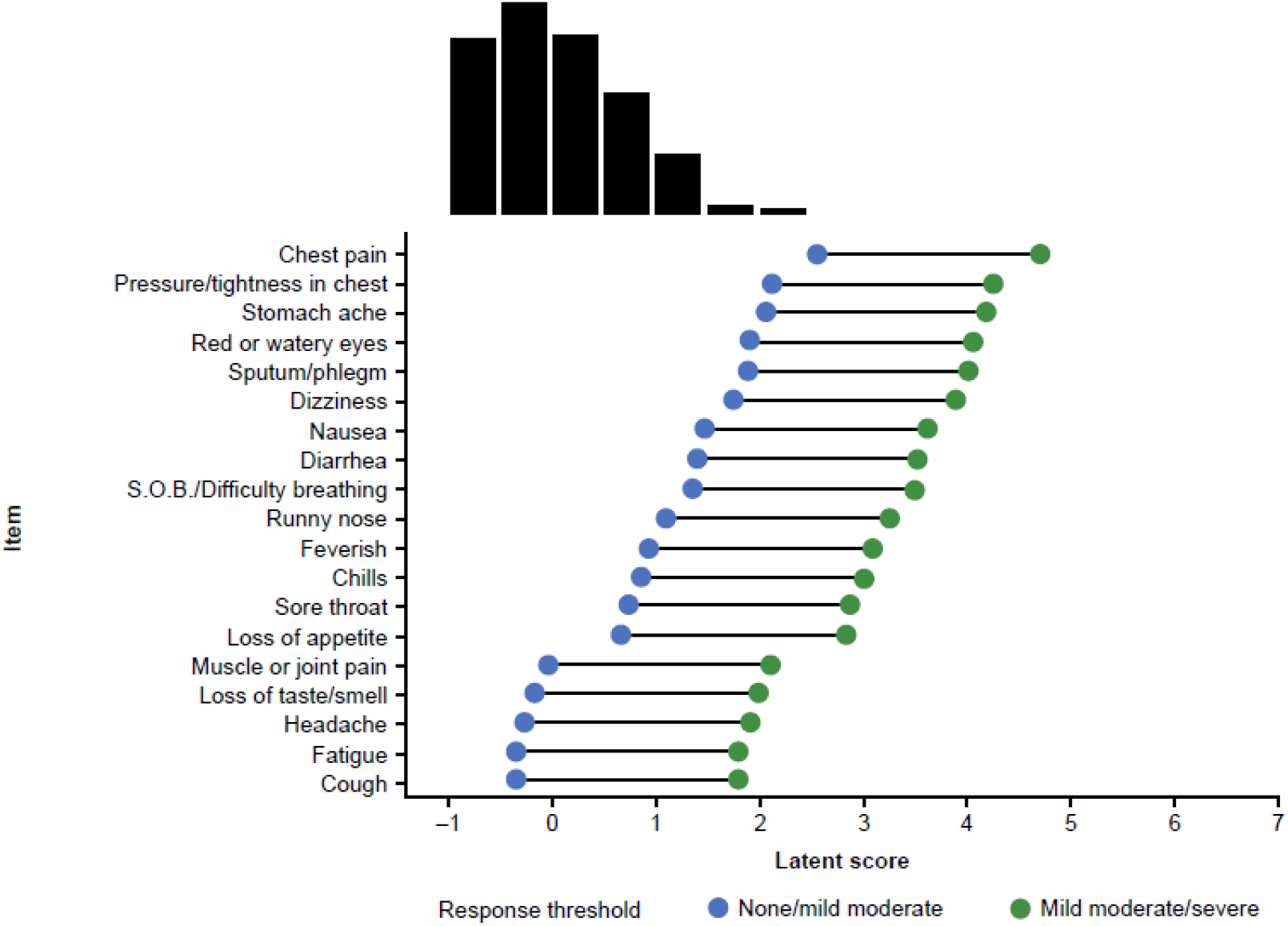
Wright map for SE-C19 items at baseline, 19 items, 3-rating scale. SE-C19=Symptoms Evolution of COVID-19; S.O.B.=shortness of breath.

**Figure 4:**
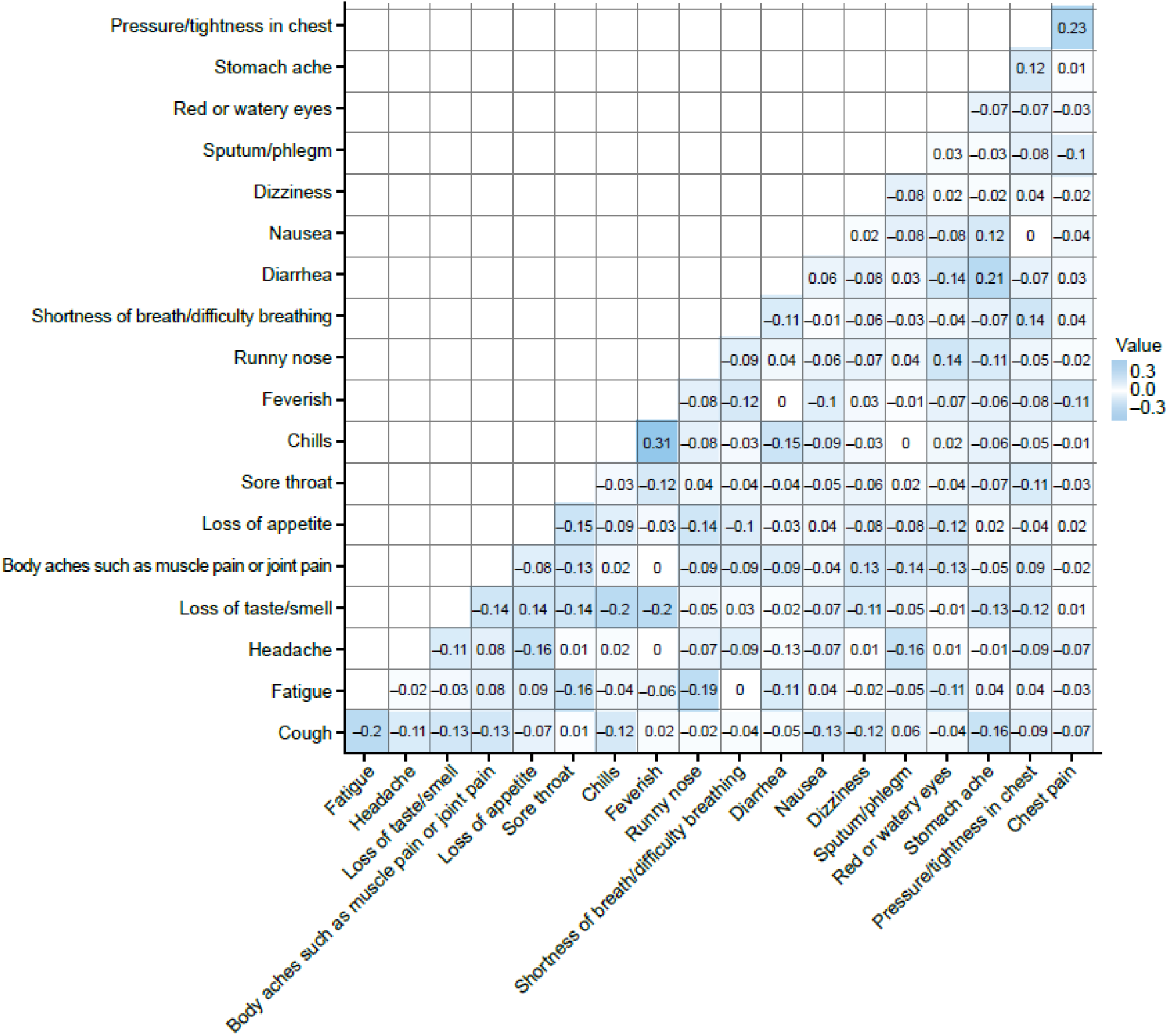
Heat map of the residual correlation matrix.

The RMT analysis confirmed overall good fit for the items, with the exception of “Vomiting”, “Confusion”, and “Rash”. “Loss of taste and smell” also showed slightly poorer model fit than expected, but it has a strong theoretical association with COVID-19 symptomology in this study population (Table S7 and Table S8).

### Development of a symptom-resolution endpoint

A symptom-resolution endpoint was developed. Based on the RMT model findings, “Vomiting”, “Confusion” and “Rash” were excluded together with “Sneezing” from the responder definition. A responder can be defined as a patient with resolution (ie, rating of *None*) of the 19 remaining items, except “Cough”, “Fatigue” and “Headache” for which a maximum of *Mild* or *Moderate* could be rated for each of these items.

## Discussion

This study, based on 657 symptomatic outpatients with COVID-19 confirmed by RT-qPCR, demonstrates that the SE-C19 is a valid and reliable measure able to capture longitudinal changes in the occurrence and severity of COVID-19-related symptoms in outpatients. To our knowledge, this is the first study to establish the reliability and validity of a patient-reported COVID-19 symptom measure for outpatients who currently represent >80% of all patients with COVID-19,^4,26^ and may become more prevalent as vaccination against COVID-19 increases and severe cases decrease.

In our study and in line with the literature,^7–11^ patients had heterogenous symptom presentations at baseline. The most prevalent symptoms among outpatients were cough, fatigue, headache, body aches and loss of taste/smell. However, each of these symptoms was only experienced by 47·0%–69·9% of patients at baseline. This heterogeneity and the most prevalent symptoms in our study are consistent with the findings of others for outpatients with COVID-19.^1,7–11^ In the current study, however, patients were more likely to report a *Moderate* or *Severe* “Loss of taste and smell” as compared to other symptoms. This was not the case in a US study with COVID-19 positive outpatients in which 4% of “Loss of taste and smell” cases were classified as *Severe* (versus 43% in our study). This difference may be due to the primary respondents being physicians rather than patients in the US study and its retrospective nature.^11^ Patient and physician perspectives are known to differ when reporting on disease severity.^27^

In line with published data from other longitudinal studies with outpatients, symptoms progression was also heterogeneous.^1,8–11^ Some symptoms such as “Feverish”, “Vomiting”, “Confusion”, and “Rash” resolved within the first week from randomisation. Other symptoms, such as “Chills”, “Nausea”, “Diarrhoea”, “Red or watery eyes”, “Dizziness”, “Pressure/tightness in chest”, and “Stomach ache” persisted generally until the end of the second week. Over the course of 3 weeks, most symptoms resolved except for “Cough”, “Headache”, and “Fatigue”. In other longitudinal clinical and observational studies, chills,^1,8,9^ fever,^1,7,8–10^ diarrhoea,^1,8,10^ and vomiting^1,9^ also resolved quickly, while weakness and fatigue,^1,8–10^ cough,^1,10^ and headache^1,8^ were the least likely symptoms to have resolved within 3 weeks. Longitudinal changes may differ across patients and COVID-19 variants.

Our analyses suggest that the SE-C19 instrument is more appropriately scored with a three-point scale (*None, Mild/Moderate*, and *Severe*) because of an apparent lack of discrimination between the *Mild* and *Moderate* response options when using RMT. Collapsing the *Mild* and *Moderate* response options simplified the rating scale structure and still allowed distinguishing severe symptom ratings from less severe symptom ratings and absence of symptoms.

At baseline, almost all patients (98%) in this population reported at least one of the 23 symptoms included in the SE-C19 questionnaire. Despite the heterogeneity of the symptom progression, the percentage of symptomatic patients gradually decreased and by end of study (ie, Day 29) most (74.4%) but not all patients were asymptomatic regardless of treatment assignment. Different symptoms resolve at different timepoints.^1,8–10^ In the context of clinical trials, it is important to use a responder definition that allows assessment of treatment benefit.^5^ The responder endpoint defined using SE-C19 is based on symptoms resolution, a commonly used endpoint in clinical trials of infectious diseases. Based on the psychometric analyses conducted in COVID-19 positive outpatients, the proposed responder definition excludes the SE-C19 item “Sneezing” because of its low ability to discriminate severity levels, and “Confusion”, “Rash”, and “Vomiting”, because of their low prevalence in this population. This is consistent with the symptomology described in other publications.^1,8–10^ In addition, the symptom resolution endpoint does not require full resolution of “Cough”, “Fatigue” and “Headache”. Patients were considered responders if they reported a maximum of *Mild* or *Moderate* on each of these three items which resolved less quickly than the others. This definition of symptoms resolution is in line with the FDA guidance for COVID-19 in outpatients,^5^ which recommends that the PRO endpoint should focus on the severity of individual item scores appraised in concert and not on aggregate scale scores. Our responder definition allows for patients to be categorised as responders or non-responders and thus, be applied in response rates and time to response endpoints in clinical trials.

For the symptom-resolution endpoint to be relevant, it is important to ensure that patients entering the trial with low symptom severity do not automatically qualify as responders. This can be done by estimating a total score based on the sum of the score (0 for *None*, 1 for *Mild/Moderate* and 2 for *Severe*) assigned to each item and including only patients with a total score >3 at baseline. The meaningfulness of this endpoint needs to be further assessed in additional studies however the findings in our study are promising and suggest that the SE-C19 and related endpoint could be used in clinical studies and, potentially, also in clinical practice.

In conclusion, we developed a symptom resolution definition based on 19 SE-C19 items that could be used as a standard in future clinical trials. The SE-C19 is easy to complete^15^ and could also be leveraged for daily symptom monitoring and decision-making in clinical practice. This study provides evidence that the SE-C19 instrument is a valid, reliable, and sensitive measure that can be used to assess COVID-19 symptoms and their progression among outpatients. Further studies are needed to assess the sensitivity and specificity of SE-C19 as a screening tool for outpatients with COVID-19. Additional studies using clinical markers or measures may also help to further understand the responder definition developed for clinical trials and its application in routine clinical practice, and to better understand the relevance of the other 4 items in monitoring the disease. Although the SE-C19 was initially designed for the outpatient setting, its potential use in hospitalised patients should be assessed. SE-C19 could be used either as additional criteria for discharge decisions, or for follow-up for potential readmission which may occur to up to 20% of patients.^28^ In sum, the SE-C19 instrument is a useful tool for tracking symptom trajectories in studies of COVID-19 outpatients and may ultimately have important applications in clinical practice.

## Supporting information

AJP ICMJE

AMR ICMJE

CG ICMJE

DR ICMJE

GF ICMJE

MH ICMJE

PG ICMJE

SS ICMJE

VM ICMJE

## Data Availability

Not applicable

## Contributors

DR contributed to conceptualisation, funding acquisition, design, methodology, project administration, supervision, visualisation, interpretation and writing (original draft, review and editing). PG, GF, and AMR contributed to the design, psychometric analysis, interpretation and writing (original draft, review and editing). CG contributed to data analysis, interpretation, and writing (original draft, review and editing). MH, SS, AP, and VM contributed to the interpretation and writing (review and editing).

## Data sharing

Not applicable.

## Acknowledgments

We thank the study participants, their families and the clinicians involved in this trial. Medical writing assistance was provided by Hervé Besson, PhD, and Montse Casamayor, MD, PhD, from IQVIA and editorial support was provided by Prime, Knutsford, United Kingdom, funded by Regeneron Pharmaceuticals, Inc.

