## Supplement for "A reliable and valid measure of COVID-19 patient-reported symptoms in outpatients: the Symptoms Evolution of COVID-19 (SE-C19) instrument"

### Supplementary material

***Table* S1: Completion rates of SE-C19, PGIS and PGIC**

|  | **SE-C19** | **PGIS** | **PGIC** |
| --- | --- | --- | --- |
| Baseline | 543/649 (83·7%) | 451/649 (69·5%) | NA |
| Day 2 | 519/647 (80·2%) | 513/647 (79·3%) | NA |
| Day 3 | 520/647 (80·4%) | 516/647 (79·8%) | NA |
| Day 4 | 509/646 (78·8%) | 504/646 (78·0%) | NA |
| Day 5 | 516/646 (79·9%) | 510/646 (78·9%) | NA |
| Day 6 | 495/646 (76·6%) | 491/646 (76·0%) | NA |
| Day 7 | 519/646 (80·3%) | 515/646 (79·7%) | NA |
| Day 8 | 506/645 (78·4%) | 502/645 (77·8%) | NA |
| Day 9 | 491/645 (76·1%) | 487/645 (75·5%) | NA |
| Day 10 | 485/644 (75·3%) | 480/644 (74·5%) | NA |
| Day 11 | 490/644 (76·1%) | 487/644 (75·6%) | NA |
| Day 12 | 489/643 (76·0%) | 484/643 (75·3%) | NA |
| Day 13 | 490/642 (76·3%) | 488/642 (76·0%) | NA |
| Day 14 | 458/641 (71·5%) | 455/641 (71·0%) | NA |
| Day 15 | 462/641 (72·1%) | 456/641 (71·1%) | NA |
| Day 16 | 481/641 (75·0%) | 481/641 (75·0%) | NA |
| Day 17 | 468/640 (73·1%) | 462/640 (72·2%) | NA |
| Day 18 | 476/637 (74·7%) | 469/637 (73·6%) | NA |
| Day 19 | 463/637 (72·7%) | 458/637 (71·9%) | NA |
| Day 20 | 473/637 (74·3%) | 468/637 (73·5%) | NA |
| Day 21 | 444/637 (69·7%) | 438/637 (68·8%) | NA |
| Day 22 | 488/637 (76·6%) | 487/637 (76·5%) | NA |
| Day 23 | 305/637 (47·9%) | 302/637 (47·4%) | NA |
| Day 24 | 284/636 (44·7%) | 284/636 (44·7%) | NA |
| Day 25 | 293/636 (46·1%) | 290/636 (45·6%) | NA |
| Day 26 | 292/634 (46·1%) | 283/634 (44·6%) | NA |
| Day 27 | 280/629 (44·5%) | 277/629 (44·0%) | NA |
| Day 28 | 253/595 (42·5%) | 250/595 (42·0%) | NA |
| Day 29 | 320/508 (63·0%) | 274/508 (53·9%) | 274/508 (53.9%) |

Completion rates are provided as patients completing the questionnaire divided by the number of patients expected to complete it at that timepoint.

PGIC=Patient Global Impression of Change; PGIS=Patient Global Impression of Severity; SE-C19=Symptoms Evolution of COVID-19.

*Table* S2. Percentages of patients with reporting no symptoms (None) at baseline, and Days 7, 15, 22, and 29

| SE-C19 item | Baseline | Day 7 | Day 15 | Day 22 | Day 29 |
| --- | --- | --- | --- | --- | --- |
| Item 1: Feverish | 68·0% | 95·2% | 97·8% | 98·8% | 99·4% |
| Item 2: Sore throat | 62·2% | 88·2% | 93·1% | 97·3% | 97·8% |
| Item 3: Cough | 30·9% | 56·1% | 77·7% | 85·9% | 90·0% |
| Item 4: Shortness of breath/Difficulty breathing | 75·3% | 87·5% | 94·4% | 94·9% | 96·9% |
| Item 5: Chills | 65·7% | 94·8% | 98·1% | 99·6% | 99·7% |
| Item 6: Nausea | 77·2% | 91·3% | 96·8% | 99·0% | 99·7% |
| Item 7: Vomiting | 95·2% | 99·2% | 99·8% | 100·0% | 100·0% |
| Item 8: Diarrhoea | 77·0% | 88·4% | 95·7% | 97·3% | 98·8% |
| Item 9: Headache | 39·4% | 66·9% | 82·0% | 89·3% | 95·0% |
| Item 10: Red or watery eyes | 83·4% | 94·8% | 97·0% | 98·4% | 99·4% |
| Item 11: Body aches (muscle pain/joint pain) | 48·1% | 82·7% | 90·3% | 94·3% | 95·9% |
| Item 12: Loss of taste/smell | 53·0% | 62·2% | 82·7% | 87·7% | 90·3% |
| Item 13: Fatigue | 38·9% | 62·8% | 80·5% | 87·1% | 91·9% |
| Item 14: Loss of appetite | 64·3% | 82·9% | 93·9% | 97·5% | 99·1% |
| Item 15: Confusion | 94·1% | 98·1% | 98·9% | 99·2% | 99·4% |
| Item 16: Dizziness | 81·2% | 92·3% | 96·8% | 98·4% | 100·0% |
| Item 17: Pressure/Tightness in chest | 86·0% | 94·4% | 97·0% | 98·2% | 97·8% |
| Item 18: Chest pain | 90·8% | 96·7% | 98·3% | 99·2% | 99·1% |
| Item 19: Stomach ache | 86·4% | 93·6% | 97·0% | 99·0% | 99·7% |
| Item 20: Rash | 97·4% | 98·7% | 99·6% | 99·4% | 99·7% |
| Item 21: Sneezing | 76·4% | 90·2% | 94.8% | 96·5% | 98·4% |
| Item 22: Runny nose | 70·0% | 88·1% | 93·3% | 96·1% | 96·6% |
| Item 23: Sputum/Phlegm | 82·5% | 86·5% | 92·0% | 94·9% | 97·2% |

***Table S3.* Intra-class correlation coefficients**

| **Symptoms** | **Day 16-17** | **Day 17-18** | **Day 18-19** | **Day 19-20** | **Day 20-21** | **Day 21-22** |
| --- | --- | --- | --- | --- | --- | --- |
| Item 1: Feverish | 0·663 | 0·496 | 0·613 | 0·499 | 0·799 | 0·665 |
| Item 2: Sore throat | 0·683 | 0·759 | 0·702 | 0·681 | 0·869 | 0·768 |
| Item 3: Cough | 0·834 | 0·882 | 0·790 | 0·847 | 0·885 | 0·878 |
| Item 4: Shortness of breath | 0·791 | 0·714 | 0·726 | 0·823 | 0·674 | 0·831 |
| Item 5: Chills | 0·798 | 0·737 | 0·699 | 0·888 | 0·889 | 0·499 |
| Item 6: Nausea | 0·566 | 0·495 | 0·245 | 0·231 | 0·868 | 0·497 |
| Item 7: Vomiting | NC | NC | NC | NC | NC | NC |
| Item 8: Diarrhoea | 0·621 | 0·643 | 0·545 | 0·530 | 0·680 | 0·774 |
| Item 9: Headache | 0·755 | 0·849 | 0·855 | 0·832 | 0·795 | 0·795 |
| Item 10: Red or watery eyes | 0·662 | 0·293 | 0·734 | 0·194 | 0·568 | 0·241 |
| Item 11: Body aches | 0·579 | 0·822 | 0·826 | 0·731 | 0·735 | 0·688 |
| Item 12: Loss of taste/smell | 0·913 | 0·916 | 0·895 | 0·974 | 0·932 | 0·929 |
| Item 13: Fatigue | 0·830 | 0·794 | 0·843 | 0·800 | 0·786 | 0·714 |
| Item 14: Loss of appetite | 0·769 | 0·757 | 0·681 | 0·670 | 0·503 | 0·596 |
| Item 15: Confusion | 0·566 | 0·415 | 0·360 | 0·666 | NC | 0·181 |
| Item 16: Dizziness | 0·689 | 0·433 | 0·641 | 0·623 | 0·321 | 0·492 |
| Item 17: Pressure in chest | 0·678 | 0·515 | 0·797 | 0·414 | 0·689 | 0·756 |
| Item 18: Chest pain | 0·857 | 0·570 | 0·598 | NC | 0·956 | 0·767 |
| Item 19: Stomach ache | 0·326 | 0·470 | 0·466 | 0·623 | 0·822 | 0·735 |
| Item 20: Rash | NC | 0·665 | 0·888 | 0·923 | 0·665 | 0·889 |
| Item 21: Sneezing | 0·612 | 0·657 | 0·700 | 0·621 | 0·764 | 0·783 |
| Item 22: Runny nose | 0·633 | 0·650 | 0·726 | 0·750 | 0·560 | 0·591 |
| Item 23: Sputum/Phlegm | 0·705 | 0·747 | 0·817 | 0·813 | 0·814 | 0·823 |

NC=not calculated.

***Table* S4. Mean difference in SE-C19 item scores at baseline between known groups defined by PGIS value at baseline (convergence validity)**

| **Item scores** | **PGIS categories** | **N** | **Mean (SD)** | **95% CI** | **P-value** |
| --- | --- | --- | --- | --- | --- |
| Item 1: Feverish | None or Mild | 208 | 0·25 (0·56) | [0·169; 0·321] | <0·0001 |
|  | Moderate or Severe | 243 | 0·73 (0·95) | [0·612; 0·853] |  |
| Item 2: Sore throat | None or Mild | 208 | 0·37 (0·61) | [0·287; 0·453] | <0·0001 |
|  | Moderate or Severe | 243 | 0·76 (0·95) | [0·637; 0·877] |  |
| Item 3: Cough | None or Mild | 208 | 0·72 (0·68) | [0·623; 0·810] | <0·0001 |
|  | Moderate or Severe | 243 | 1·34 (0·92) | [1·225; 1·458] |  |
| Item 4: Shortness of breath | None or Mild | 208 | 0·20 (0·47) | [0·133; 0·261] | <0·0001 |
|  | Moderate or Severe | 243 | 0·55 (0·89) | [0·439; 0·663] |  |
| Item 5: Chills | None or Mild | 208 | 0·32 (0·62) | [0·237; 0·407] | <0·0001 |
|  | Moderate or Severe | 243 | 0·74 (0·98) | [0·617; 0·864] |  |
| Item 6: Nausea | None or Mild | 208 | 0·15 (0·45) | [0·093; 0·215] | <0·0001 |
|  | Moderate or Severe | 243 | 0·49 (0·83) | [0·390; 0·598] |  |
| Item 7: Vomiting | None or Mild | 208 | 0·03 (0·22) | [-0·001; 0·059] | 0·017 |
|  | Moderate or Severe | 243 | 0·10 (0·40) | [0·048; 0·150] |  |
| Item 8: Diarrhoea | None or Mild | 208 | 0·19 (0·51) | [0·118; 0·257] | <0·0001 |
|  | Moderate or Severe | 243 | 0·56 (0·92) | [0·443; 0·676] |  |
| Item 9: Headache | None or Mild | 208 | 0·66 (0·78) | [0·552; 0·766] | <0·0001 |
|  | Moderate or Severe | 243 | 1·42 (1·07) | [1·289; 1·559] |  |
| Item 10: Red or watery eyes | None or Mild | 208 | 0·19 (0·46) | [0·125; 0·250] | 0·392 |
|  | Moderate or Severe | 243 | 0·28 (0·64) | [0·194; 0·357] |  |
| Item 11: Body aches | None or Mild | 208 | 0·54 (0·80) | [0·434; 0·652] | <0·0001 |
|  | Moderate or Severe | 243 | 1·35 (1·18) | [1·205; 1·503] |  |
| Item 12: Loss of taste/smell | None or Mild | 208 | 0·71 (1·04) | [0·569; 0·854] | <0·0001 |
|  | Moderate or Severe | 243 | 1·38 (1·31) | [1·218; 1·548] |  |
| Item 13: Fatigue | None or Mild | 208 | 0·82 (0·89) | [0·696; 0·939] | <0·0001 |
|  | Moderate or Severe | 243 | 1·54 (1·12) | [1·398; 1·680] |  |
| Item 14: Loss of appetite | None or Mild | 208 | 0·37 (0·70) | [0·270; 0·461] | <0·0001 |
|  | Moderate or Severe | 243 | 0·95 (1·13) | [0·812; 1·097] |  |
| Item 15: Confusion | None or Mild | 208 | 0·02 (0·14) | [0·000; 0·038] | 0·002 |
|  | Moderate or Severe | 243 | 0·16 (0·55) | [0·087; 0·225] |  |
| Item 16: Dizziness | None or Mild | 208 | 0·09 (0·34) | [0·046; 0·137] | <0·0001 |
|  | Moderate or Severe | 243 | 0·47 (0·80) | [0·372; 0·575] |  |
| Item 17: Pressure in chest | None or Mild | 208 | 0·12 (0·38) | [0·064; 0·167] | 0·009 |
|  | Moderate or Severe | 243 | 0·28 (0·66) | [0·197; 0·363] |  |
| Item 18: Chest pain | None or Mild | 208 | 0·07 (0·29) | [0·028; 0·107] | 0·006 |
|  | Moderate or Severe | 243 | 0·23 (0·63) | [0·147; 0·306] |  |
| Item 19: Stomach ache | None or Mild | 208 | 0·09 (0·35) | [0·044; 0·139] | <0·0001 |
|  | Moderate or Severe | 243 | 0·33 (0·73) | [0·237; 0·422] |  |
| Item 20: Rash | None or Mild | 208 | 0·02 (0·17) | [-0·004; 0·042] | 0·203 |
|  | Moderate or Severe | 243 | 0·06 (0·35) | [0·014; 0·102] |  |
| Item 21: Sneezing | None or Mild | 208 | 0·22 (0·47) | [0·157; 0·285] | 0·006 |
|  | Moderate or Severe | 243 | 0·42 (0·70) | [0·327; 0·504] |  |
| Item 22: Runny nose | None or Mild | 208 | 0·34 (0·58) | [0·257; 0·416] | 0·055 |
|  | Moderate or Severe | 243 | 0·51 (0·80) | [0·413; 0·616] |  |
| Item 23: Sputum/Phlegm | None or Mild | 208 | 0·18 (0·44) | [0·117; 0·238] | 0·036 |
|  | Moderate or Severe | 243 | 0·35 (0·70) | [0·257; 0·434] |  |

CI=confidence interval; PGIS=Patient Global Impression of Severity; SD=standard deviation.

***Table S5: Mean change in SE-C19 item scores from baseline to Days 15 and 29 by PGIS scores at Days 15 and 29, respectively***

|  |  | **At Day 15** | | | | **At Day 29** | | | |
| --- | --- | --- | --- | --- | --- | --- | --- | --- | --- |
| **Item scores** | **PGIS group** | **N** | **Mean (SD)** | **Median [95% CI]** | **P-value** | **N** | **Mean (SD)** | **Median [95% CI]** | **P-value** |
| Item 1: Feverish | Any improvement | 222 | -0·49 (0·82) | 0 [-0·600; -0·382] | 0·167 | 184 | -0·46 (0·75) | 0 [-0·566; -0·347] | 0·247 |
|  | No change or worsening | 114 | -0·35 (0·76) | 0 [-0·493; -0·209] |  | 33 | -0·24 (0·71) | 0 [-0·494; 0·009] |  |
| Item 2: Sore throat | Any improvement | 222 | -0·53 (0·83) | 0 [-0·636; -0·418] | 0·178 | 184 | -0·52 (0·80) | 0 [-0·637; -0·406] | 0·017 |
|  | No change or worsening | 114 | -0·39 (0·76) | 0 [-0·527; -0·245] |  | 33 | -0·18 (0·64) | 0 [-0·407; 0·043] |  |
| Item 3: Cough | Any improvement | 222 | -0·94 (0·86) | -1 [-1·051; -0·823] | <0·0001 | 184 | -1·04 (0·77) | -1 [-1·156; -0·931] | 0·005 |
|  | No change or worsening | 114 | -0·46 (0·86) | 0 [-0·625; -0·305] |  | 33 | -0·61 (0·83) | 0 [-0·899; -0·313] |  |
| Item 4: Shortness of breath | Any improvement | 222 | -0·33 (0·72) | 0 [-0·428; -0·239] | 0·052 | 184 | -0·34 (0·71) | 0 [-0·445; -0·240] | 0·439 |
|  | No change or worsening | 114 | -0·14 (0·61) | 0 [-0·253; -0·028] |  | 33 | -0·15 (0·83) | 0 [-0·447; 0·144] |  |
| Item 5: Chills | Any improvement | 222 | -0·54 (0·91) | 0 [-0·656; -0·416] | 0·127 | 184 | -0·54 (0·82) | 0 [-0·657; -0·419] | 0·100 |
|  | No change or worsening | 114 | -0·35 (0·66) | 0 [-0·474; -0·227] |  | 33 | -0·27 (0·57) | 0 [-0·476; -0·069] |  |
| Item 6: Nausea | Any improvement | 222 | -0·31 (0·68) | 0 [-0·400; -0·221] | 0·285 | 184 | -0·32 (0·70) | 0 [-0·423; -0·219] | 0·907 |
|  | No change or worsening | 114 | -0·20 (0·67) | 0 [-0·326; -0·078] |  | 33 | -0·30 (0·64) | 0 [-0·529; -0·077] |  |
| Item 7: Vomiting | Any improvement | 222 | -0·07 (0·39) | 0 [-0·123; -0·021] | 0·939 | 184 | -0·06 (0·33) | 0 [-0·108; -0·011] | 0·554 |
|  | No change or worsening | 114 | -0·04 (0·21) | 0 [-0·082; -0·006] |  | 33 | -0·09 (0·38) | 0 [-0·227; 0·045] |  |
| Item 8: Diarrhoea | Any improvement | 222 | -0·36 (0·80) | 0 [-0·462; -0·250] | 0·973 | 184 | -0·36 (0·76) | 0 [-0·474; -0·254] | 0·352 |
|  | No change or worsening | 114 | -0·25 (0·73) | 0 [-0·389; -0·120] |  | 33 | -0·24 (0·66) | 0 [-0·477; -0·007] |  |
| Item 9: Headache | Any improvement | 222 | -1·06 (1·05) | -1 [-1·202; -0·924] | <0·0001 | 184 | -1·08 (0·97) | -1 [-1·222; -0·941] | 0·001 |
|  | No change or worsening | 114 | -0·33 (0·95) | 0 [-0·509; -0·158] |  | 33 | -0·45 (0·87) | 0 [-0·763; -0·146] |  |
| Item 10: Red or watery eyes | Any improvement | 222 | -0·19 (0·56) | 0 [-0·264; -0·115] | 0·299 | 184 | -0·22 (0·54) | 0 [-0·302; -0·144] | 0·798 |
|  | No change or worsening | 114 | -0·22 (0·77) | 0 [-0·363; -0·076] |  | 33 | -0·21 (0·60) | 0 [-0·425; 0·001] |  |
| Item 11: Body aches | Any improvement | 222 | -0·99 (1·14) | -1 [-1·137; -0·836] | 0·001 | 184 | -0·95 (1·03) | -1 [-1·095; -0·796] | 0·155 |
|  | No change or worsening | 114 | -0·53 (0·93) | 0 [-0·699; -0·353] |  | 33 | -0·64 (1·06) | 0 [-1·011; -0·262] |  |
| Item 12: Loss of taste/smell | Any improvement | 222 | -0·96 (1·30) | 0 [-1·132; -0·787] | 0·002 | 184 | -0·99 (1·23) | 0 [-1·168; -0·810] | 0·099 |
|  | No change or worsening | 114 | -0·46 (1·17) | 0 [-0·673; -0·239] |  | 33 | -0·58 (1·20) | 0 [-1·001; -0·150] |  |
| Item 13: Fatigue | Any improvement | 222 | -1·11 (1·06) | -1 [-1·253; -0·972] | <0·0001 | 184 | -1·20 (1·07) | -1 [-1·357; -1·045] | 0·063 |
|  | No change or worsening | 114 | -0·58 (1·00) | 0 [-0·765; -0·393] |  | 33 | -0·79 (1·19) | 0 [-1·211; -0·365] |  |
| Item 14: Loss of appetite | Any improvement | 222 | -0·68 (1·05) | 0 [-0·824; -0·545] | 0·201 | 184 | -0·66 (1·00) | 0 [-0·809; -0·518] | 0·574 |
|  | No change or worsening | 114 | -0·47 (0·91) | 0 [-0·643; -0·304] |  | 33 | -0·52 (0·83) | 0 [-0·811; -0·220] |  |
| Item 15: Confusion | Any improvement | 222 | -0·08 (0·37) | 0 [-0·130; -0·032] | 0·284 | 184 | -0·09 (0·40) | 0 [-0·144; -0·029] | 0·619 |
|  | No change or worsening | 114 | -0·04 (0·31) | 0 [-0·101; 0·013] |  | 33 | -0·06 (0·43) | 0 [-0·213; 0·091] |  |
| Item 16: Dizziness | Any improvement | 222 | -0·31 (0·72) | 0 [-0·406; -0·216] | 0·206 | 184 | -0·27 (0·62) | 0 [-0·356; -0·176] | 0·623 |
|  | No change or worsening | 114 | -0·20 (0·58) | 0 [-0·310; -0·094] |  | 33 | -0·33 (0·69) | 0 [-0·579; -0·088] |  |
| Item 17: Pressure in chest | Any improvement | 222 | -0·18 (0·48) | 0 [-0·239; -0·113] | 0·017 | 184 | -0·17 (0·49) | 0 [-0·246; -0·102] | 0·21 |
|  | No change or worsening | 114 | -0·04 (0·53) | 0 [-0·134; 0·063] |  | 33 | -0·06 (1·00) | 0 [-0·415; 0·293] |  |
| Item 18: Chest pain | Any improvement | 222 | -0·15 (0·47) | 0 [-0·210; -0·087] | 0·033 | 184 | -0·11 (0·42) | 0 [-0·169; -0·048] | 0·226 |
|  | No change or worsening | 114 | -0·07 (0·65) | 0 [-0·190; 0·050] |  | 33 | -0·06 (0·90) | 0 [-0·379; 0·258] |  |
| Item 19: Stomach ache | Any improvement | 222 | -0·18 (0·53) | 0 [-0·246; -0·106] | 0·414 | 184 | -0·20 (0·56) | 0 [-0·283; -0·120] | 0·514 |
|  | No change or worsening | 114 | -0·11 (0·65) | 0 [-0·234; 0·006] |  | 33 | -0·24 (0·56) | 0 [-0·441; -0·044] |  |
| Item 20: Rash | Any improvement | 222 | -0·05 (0·32) | 0 [-0·092; -0·007] | 0·966 | 184 | -0·04 (0·35) | 0 [-0·089; 0·013] | 0·296 |
|  | No change or worsening | 114 | -0·04 (0·31) | 0 [-0·101; 0·013] |  | 33 | -0·06 (0·24) | 0 [-0·147; 0·025] |  |
| Item 21: Sneezing | Any improvement | 222 | -0·31 (0·62) | 0 [-0·393; -0·229] | 0·096 | 184 | -0·34 (0·60) | 0 [-0·429; -0·255] | 0·320 |
|  | No change or worsening | 114 | -0·21 (0·71) | 0 [-0·342; -0·079] |  | 33 | -0·21 (0·60) | 0 [-0·425; 0·001] |  |
| Item 22: Runny nose | Any improvement | 222 | -0·40 (0·70) | 0 [-0·488; -0·304] | 0·039 | 184 | -0·49 (0·71) | 0 [-0·598; -0·391] | 0·044 |
|  | No change or worsening | 114 | -0·18 (0·70) | 0 [-0·314; -0·055] |  | 33 | -0·18 (0·58) | 0 [-0·389; 0·025] |  |
| Item 23: Sputum/Phlegm | Any improvement | 222 | -0·17 (0·54) | 0 [-0·243; -0·099] | 0·894 | 184 | -0·28 (0·60) | 0 [-0·369; -0·196] | 0·722 |
|  | No change or worsening | 114 | -0·19 (0·62) | 0 [-0·308; -0·078] |  | 33 | -0·36 (0·82) | 0 [-0·655; -0·072] |  |

P-value based on Mann-Whitney U test, also known as Wilcoxon Rank Sum test, a nonparametric alternative to an independent sample t-test.

Any improvement group includes those patients whose PGIS scores at Day 15 or Day 29 improve at least one category from baseline (i.e.: from Mild to None). No change or worsening group includes those patients with the same PGIS score at Day 15 or Day 29 and baseline or those patients whose PGIS scores at Day 15 or Day 29 worsened at least 1 score from baseline (i.e.: from Mild to Moderate).

CI=confidence interval; N=Number of patients; SD=standard deviation.

***Table* S6: Mean change in SE-C19 item scores from baseline to Day 29 by PGIC scores at Day 29**

| **Item scores** | **PGIC group** | **N** | **Mean (SD)** | **Median [95% CI]** | | **P-value** |
| --- | --- | --- | --- | --- | --- | --- |
| Item 1: Feverish | Any improvement | 234 | -0·49 (0·79) | 0 | [-0·589; -0·386] | 0·025 |
|  | No change or worsening | 11 | 0·00 (0·00) | 0 | [0·000; 0·000] |  |
| Item 2: Sore throat | Any improvement | 234 | -0·47 (0·78) | 0 | [-0·571; -0·369] | 0·75 |
|  | No change or worsening | 11 | -0·55 (0·82) | 0 | [-1·096; 0·006] |  |
| Item 3: Cough | Any improvement | 234 | -0·96 (0·77) | -1 | [-1·061; -0·862] | 0·252 |
|  | No change or worsening | 11 | -0·82 (1·25) | 0 | [-1·658; 0·022] |  |
| Item 4: Shortness of breath | Any improvement | 234 | -0·31 (0·68) | 0 | [-0·399; -0·225] | 0·986 |
|  | No change or worsening | 11 | -0·36 (1·29) | 0 | [-1·228; 0·501] |  |
| Item 5: Chills | Any improvement | 234 | -0·55 (0·82) | 0 | [-0·657; -0·446] | 0·066 |
|  | No change or worsening | 11 | -0·09 (0·30) | 0 | [-0·293; 0·112] |  |
| Item 6: Nausea | Any improvement | 234 | -0·33 (0·70) | 0 | [-0·423; -0·243] | 0·306 |
|  | No change or worsening | 11 | -0·09 (0·30) | 0 | [-0·293; 0·112] |  |
| Item 7: Vomiting | Any improvement | 234 | -0·06 (0·33) | 0 | [-0·102; -0·018] | 0·513 |
|  | No change or worsening | 11 | 0·00 (0·00) | 0 | [0·000; 0·000] |  |
| Item 8: Diarrhoea | Any improvement | 234 | -0·35 (0·75) | 0 | [-0·451; -0·259] | 0·357 |
|  | No change or worsening | 11 | -0·18 (0·60) | 0 | [-0·587; 0·223] |  |
| Item 9: Headache | Any improvement | 234 | -1·01 (0·98) | -1 | [-1·139; -0·887] | 0·724 |
|  | No change or worsening | 11 | -0·91 (1·04) | 0 | [-1·611; -0·207] |  |
| Item 10: Red or watery eyes | Any improvement | 234 | -0·23 (0·56) | 0 | [-0·299; -0·154] | 0·161 |
|  | No change or worsening | 11 | 0·00 (0·00) | 0 | [0·000; 0·000] |  |
| Item 11: Body aches | Any improvement | 234 | -0·94 (1·04) | -1 | [-1·070; -0·801] | 0·226 |
|  | No change or worsening | 11 | -0·55 (0·82) | 0 | [-1·096; 0·006] |  |
| Item 12: Loss of taste/smell | Any improvement | 234 | -0·89 (1·24) | 0 | [-1·053; -0·733] | 0·885 |
|  | No change or worsening | 11 | -0·73 (1·01) | 0 | [-1·405; -0·049] |  |
| Item 13: Fatigue | Any improvement | 234 | -1·18 (1·10) | -1 | [-1·321; -1·038] | 0·107 |
|  | No change or worsening | 11 | -0·55 (1·37) | 0 | [-1·465; 0·374] |  |
| Item 14: Loss of appetite | Any improvement | 234 | -0·65 (0·97) | 0 | [-0·770; -0·520] | 0·294 |
|  | No change or worsening | 11 | -0·36 (0·81) | 0 | [-0·907; 0·180] |  |
| Item 15: Confusion | Any improvement | 234 | -0·08 (0·39) | 0 | [-0·127; -0·027] | 0·57 |
|  | No change or worsening | 11 | -0·09 (0·30) | 0 | [-0·293; 0·112] |  |
| Item 16: Dizziness | Any improvement | 234 | -0·26 (0·61) | 0 | [-0·344; -0·186] | 0·395 |
|  | No change or worsening | 11 | -0·45 (0·82) | 0 | [-1·006; 0·096] |  |
| Item 17: Pressure in chest | Any improvement | 234 | -0·16 (0·54) | 0 | [-0·232; -0·093] | 0·342 |
|  | No change or worsening | 11 | -0·09 (1·14) | 0 | [-0·854; 0·672] |  |
| Item 18: Chest pain | Any improvement | 234 | -0·10 (0·50) | 0 | [-0·163; -0·034] | 0·463 |
|  | No change or worsening | 11 | 0·00 (0·00) | 0 | [0·000; 0·000] |  |
| Item 19: Stomach ache | Any improvement | 234 | -0·20 (0·55) | 0 | [-0·272; -0·129] | 0·193 |
|  | No change or worsening | 11 | 0·00 (0·00) | 0 | [0·000; 0·000] |  |
| Item 20: Rash | Any improvement | 234 | -0·05 (0·35) | 0 | [-0·092; -0·002] | 0·612 |
|  | No change or worsening | 11 | 0·00 (0·00) | 0 | [0·000; 0·000] |  |
| Item 21: Sneezing | Any improvement | 234 | -0·31 (0·59) | 0 | [-0·384; -0·231] | 0·966 |
|  | No change or worsening | 11 | -0·27 (0·47) | 0 | [-0·587; 0·041] |  |
| Item 22: Runny nose | Any improvement | 234 | -0·44 (0·70) | 0 | [-0·534; -0·354] | 0·33 |
|  | No change or worsening | 11 | -0·27 (0·65) | 0 | [-0·707; 0·162] |  |
| Item 23: Sputum/Phlegm | Any improvement | 234 | -0·27 (0·62) | 0 | [-0·353; -0·194] | 0·949 |
|  | No change or worsening | 11 | -0·27 (0·65) | 0 | [-0·707; 0·162] |  |

P-value based on Mann-Whitney U test. Any improvement group includes those patients whose PGIC scores at Day 29 improve at least 1 category from baseline (i.e.: from Mild to None). No change or worsening group includes those patients with the same PGIC score at Day 29 and baseline or those patients whose PGIC scores at Day 29 worsened at least 1 score from baseline (i.e.: from Mild to Moderate).

CI=confidence interval; N=number of patients; PGIS=Patient Global Impression of Severity; SD=standard deviation.

***Table* S7: Goodness-of-fit measures for items for each model**

| **SE-C19 Items** | **22-items* and 3-ratings** | | **19-items^#^ and 3-ratings** | |
| --- | --- | --- | --- | --- |
|  | outfit | infit | outfit | infit |
| Item 1: Feverish | 0·851 | 0·939 | 0·845 | 0·932 |
| Item 2: Sore throat | 0·985 | 0·964 | 0·978 | 0·962 |
| Item 3: Cough | 0·739 | 0·724 | 0·731 | 0·715 |
| Item 4: Shortness of breath/Difficulty breathing | 0·905 | 0·972 | 0·899 | 0·971 |
| Item 5: Chills | 0·803 | 0·872 | 0·804 | 0·875 |
| Item 6: Nausea | 0·792 | 0·936 | 0·794 | 0·939 |
| Item 7: Vomiting | 0·610 | 1·053 | N/A | N/A |
| Item 8: Diarrhoea | 0·919 | 1·065 | 0·925 | 1·070 |
| Item 9: Headache | 0·809 | 0·825 | 0·806 | 0·822 |
| Item 10: Red or watery eyes | 1·010 | 1·048 | 1·006 | 1·047 |
| Item 11: Body aches such as muscle pain or joint pain | 0·889 | 0·938 | 0·892 | 0·938 |
| Item 12: Loss of taste/smell | 1·377 | 1·442 | 1·383 | 1·449 |
| Item 13: Fatigue | 0·796 | 0·814 | 0·794 | 0·811 |
| Item 14: Loss of appetite | 0·928 | 1·029 | 0·929 | 1·031 |
| Item 15: Confusion | 0·874 | 1·176 | N/A | N/A |
| Item 16: Dizziness | 0·835 | 0·923 | 0·845 | 0·927 |
| Item 17: Pressure/Tightness in chest | 0·770 | 0·935 | 0·766 | 0·938 |
| Item 18: Chest pain | 0·699 | 1·037 | 0·697 | 1·044 |
| Item 19: Stomach ache | 0·831 | 1·072 | 0·851 | 1·082 |
| Item 20: Rash | 1·075 | 1·477 | N/A | N/A |
| Item 22: Runny nose | 0·898 | 0·951 | 0·889 | 0·944 |
| Item 23: Sputum/Phlegm | 0·923 | 0·965 | 0·919 | 0·963 |

*Excluding Sneezing. #Exclduing Sneezing, Vomiting, Confusion and Rash. The 3 ratings were None, Mild/Moderate and Severe.

***Table*** **S8: RMT model person goodness-of-fit measures**

| **RMT Type of Scale** | **Statistic** | **outfit** | **z.outfit** | **infit** | **z.infit** | **Zh** |
| --- | --- | --- | --- | --- | --- | --- |
| Three levels of severity | Average | 0·878 | -0·037 | 0·910 | -0·159 | 0·122 |
|  | Standard deviation | 0·616 | 0·925 | 0·380 | 1·033 | 0·932 |
|  | Misfitting patients | n=20 (3·76%) |  |  |  |  |
| Three levels of severity (Short-form) | Average | 0·882 | -0·124 | 0·908 | -0·171 | 0·132 |
|  | Standard deviation | 0·434 | 0·948 | 0·378 | 1·027 | 0·909 |
|  | Misfitting patients | n=20 (3·76%) |  |  |  |  |

RMT=Rasch Measurement Theory.

*Figure* S1: Inter-items correlations

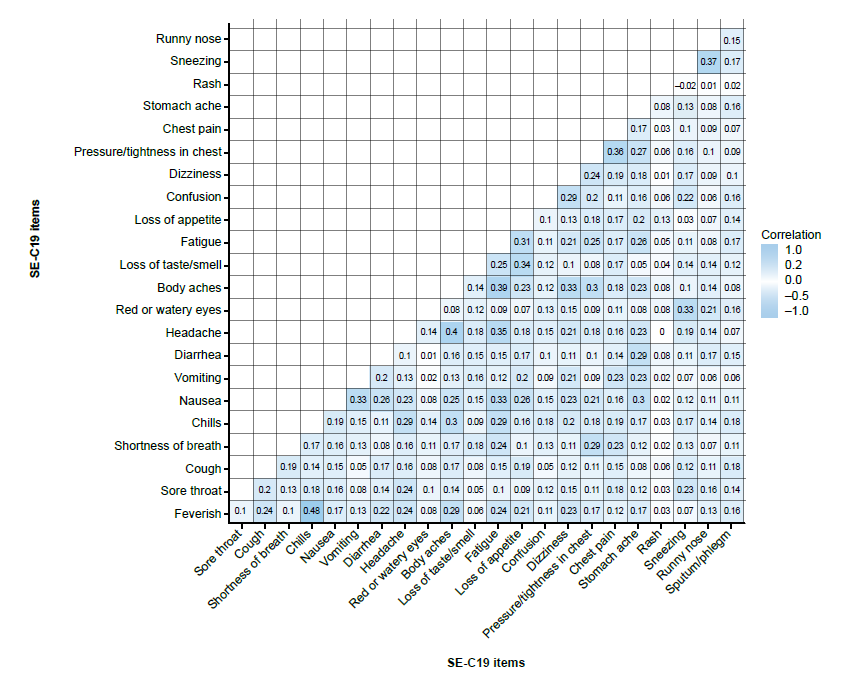

SE-C19=Symptoms Evolution of COVID-19.
